# VACS: VAccination disComfort Scale

**DOI:** 10.1101/2022.10.18.22281162

**Authors:** Manolis Wallace, Stavros Antonopoulos, Vassilis Poulopoulos

## Abstract

In this article we focus on the discomfort experienced by children during vaccination and ask ourselves how this discomfort could be quantified. We develop VACS, a tool to measure this discomfort as a number in the range 0-25 and apply it to 40 vaccinations of children aged 2 to 12. Our findings show that approximately 40% of the children do not face discomfort during vaccination, but for the rest discomfort of varying degrees is observed. We also find that doctors are content with their patients facing considerably higher discomfort levels that what the children themselves are willing to withstand. Surprisingly, characteristics such as a) gender, b) whether the state’s recommended vaccination program has been implemented in full and even c) prior negative vaccination experiences are found to be poor predictors of vaccination discomfort. Age may be a factor, with younger children experiencing discomfort more often and more intensely, but more research is required in order to validate this. The formulation of VACS opens the door for more systematic work towards the minimization of vaccination discomfort for children.

## Introduction

Vaccination is a great tool for the protection of society from many viruses that would otherwise (continue to) have a greater impact. But with needles being a source of discomfort or even fear for so many, the vaccination procedure is not pleasant for all. As immunization schedules around the world typically cover ages from birth until 18 years of age and most vaccinations take place during the first years of childhood, it is particularly young children that suffer through the vaccination procedure. But to this day we have mainly taken this discomfort for granted and forced our children to push through it, without giving any more consideration to it.

In this work we develop a new scale, VAccination disComfort Scale (VACS), that is intended to measure children’s discomfort during vaccination. VACS is based on doctor’s observations before, during and after vaccination. It adds less than one minute to the overall duration of the vaccination procedure and produces a number in the range 0-25, with 0 indicating total comfort and 25 indicating maximum discomfort. Measuring discomfort is the first step to 1) gaining a deeper understanding of who and to what extent experiences this discomfort and 2) to assessing whether different vaccination procedures might be more pleasant for children.

The remaining of this article is organized as follows: In section 2 we develop VACS and present the methodology we have followed in order to validate it. In section 3 we present the findings of our clinical trial while in section 4 we elaborate further on our findings, discuss the implications of our work and identify weakness and potential future directions.

## 2. Materials and Methods

### 2.1. VACS structure

In order to measure children’s discomfort during vaccination in as much as possible objective way, we base our calculations on the doctor’s observations. The patient is observed from the moment they enter the room or area where the vaccination takes place until they leave and a number of parameters indicating distress and discomfort are noted.

Recorded observations come from 4 distinct stages:

1. Stage I - Entrance. How the child is/behaves when entering the examination area;
2. Stage II - Examination. How the child is/behaves while being examined by the doctor and building up to the actual examination;
3. Stage III - Procedure. How the child is/behaves during the actual medical procedure of vaccination;
4. Stage IV - Completion. How the child is/behaves when the actual vaccination has been completed.

Weights are assigned to each parameter, so that a single overall value of discomfort can be computed.

### 2.2. VACS parameters

#### 2.2.1. Crying

Crying is of course a major indication of discomfort during vaccination, especially for younger children. It can be an indicator not only of pain but also of fear or other types of distress. That is why it is not only observed during/after the vaccination shot but very often even before that. The extent of crying can be varying and this can be indicative of the level of discomfort. In order to take the extent of crying into account, we use the following levels:

1. No crying;
2. Light moaning, or intermitted crying;
3. Loud crying, constant howling or sobbing.

#### 2.2.2. Hesitation

Hesitation (or the lack of it) is observed as the child enters the vaccination room/area. Hesitation is a key element that can at times even escalate to a total refusal to enter the doctor’s office. The obvious cases are:

1. No hesitation;
2. Hesitation.

#### 2.2.3. Activity

The child’s activity at various stages of the procedure is a key element that may or may not coexist with other observed parameters. Three distinct levels are considered:

1. Relaxed posture, where the child is relaxed, lying down or at another position that is normal for the child’s age and moves easily;
2. Twisting, moving back and forth, generally being in tension;
3. Assuming a defensive of fetal position, or being rigidly fully stretched.

#### 2.2.4. Facial expressions

Facial expressions are great indicators of how one feels and could not possible be left without consideration in this work. Three categories are identified as follows:

1. Relaxed muscles, smiles, showing comfort;
2. Occasional grimaces with tight facial muscles, furrowed brow, chin or jaw;
3. A continuous grimace, frequent or constant chin shaking, clenched jaw.

#### 2.2.5. Support

We observe the support that the child may need in order to calm down, as follows:

1. Child is asleep or child is awake but content and relaxed;
2. The child whines but can be calmed by touching, hugging or talking;
3. The child remains inconsolable no matter what.

#### 2.2.6. Cooperation

The way the child (eventually) cooperated for the vaccination to take place is a great indicator of the level of discomfort. We classify cooperation based on who was required to be physically involve to help (or potentially restrain) the child for the vaccination to take place, as follows:

1. The child makes it on their own;
2. The parent/guardian needs to be involved;
3. The parent/guardian and also the clinic’s staff need to be involved;
4. The parent/guardian is asked to step back, the clinic’s staff take over on their own and restrain the child as needed.

### 2.3. VACS parameter weights

The four stages that we are considering (entry, examination, procedure and completion) do not all have the same importance when discussing vaccination discomfort. It is the time leading up to vaccination and the actual act of vaccinating that cause the most discomfort to children. This is reflected in the distribution of the scores for the different stages of VACS; as can be seen in Table 1, most points are awarded for observations during the examination and procedure stages, whilst the stage when the vaccination have been completed carries the least weight.

**Table 1.** Distribution of VACS parameter weights in the four stages.

| Stage | Maximum points | Percentage |
| --- | --- | --- |
| Stage I - Entry | 5 | 20% |
| Stage II - Examination | 8 | 32% |
| Stage III – Procedure | 8 | 32% |
| Stage IV – Completion | 4 | 16% |

For all of the examined parameters, the first category listed in the previous sub-section is the one indicating no discomfort and therefore does not contribute any score to the VACS measurement. The other categories are scored as follows:

#### 2.3.1. Stage I - Entrance

Hesitation to enter the vaccination area is scored with 3 points. Light intermitted crying is scored with 1 point whilst loud constant crying contributes 2 points, reaching a maximum of 5 points for the entrance stage.

#### 2.3.2. Stage II - Examination

Occasional grimaces get 1 point and a constant grimace is scored with 2 points. Light intermitted crying is scored with 1 point whilst loud constant crying contributes 2 points. Extensive activity gets 2 points and defensive/fetal position gets 4 points, reaching a maximum of 8 points for the examination stage.

#### 2.3.3. Stage III - Procedure

Being agitated but able to calm down awards 1 point while being inconsolable awards 2 points. Requiring the assistance of a parent or guardian to complete the vaccination awards one point. 3 points are awarded if the staff need to be involved and 4 points if the parent/guardian needs to step away for the staff to take over alone. Light intermitted crying is scored with 1 point whilst loud constant crying contributes 2 points, reaching a maximum of 8 points for the procedure stage.

#### 2.3.4. Stage IV - Completion

Light intermitted crying is scored with 1 point whilst loud constant crying con-tributes 2 points. Extensive activity gets 1 point and defensive/fetal position gets 2 points, reaching a maximum of 4 points for the examination stage.

The weights are summarized in Figure 1, for easy reference.

**Figure 1.**
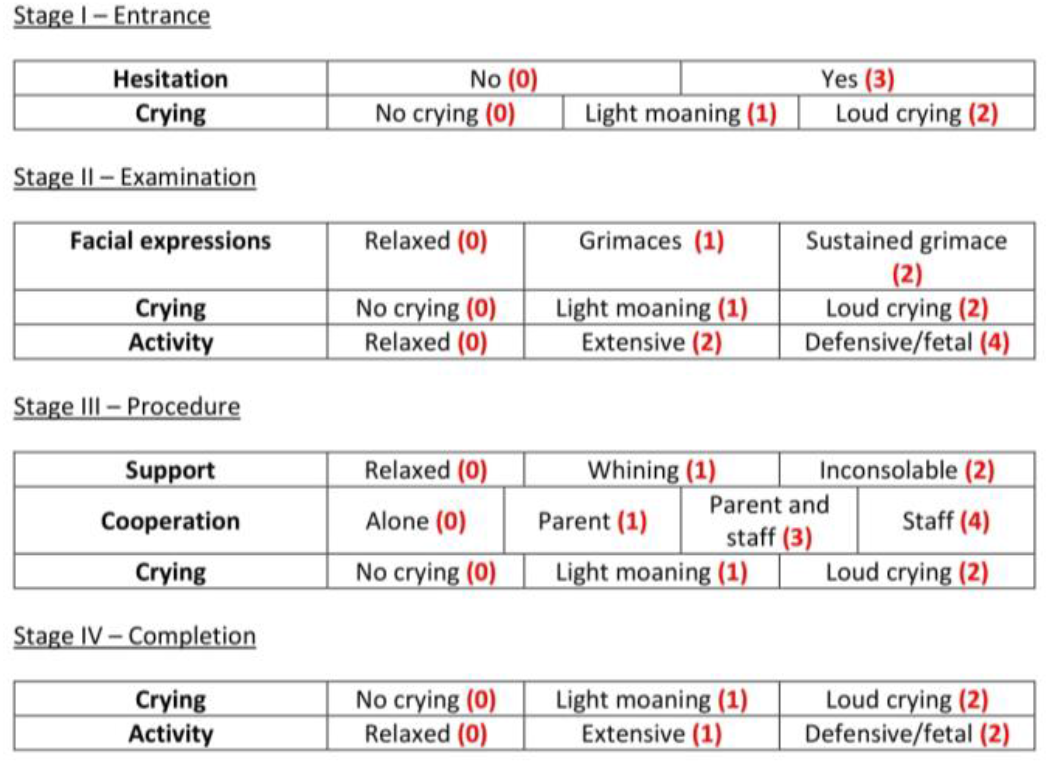
The weights of parameters considered in VACS.

### 2.4. Clinical settings

In order to validate VACS (do larger measurements truly correspond to greater discomfort?) and to set the thresholds (what is a typical VACS measurement? which measurement is excessive?) we have applied it in a clinical setting. Participants were recruited using convenience sampling. Specifically, all children visiting one of the pediatric clinics participating in this research (one public and two private ones) were considered. The two inclusion criteria were

1. The child being between 2 and 12 years one;
2. The accompanying parent/guardian being sufficiently fluent in Greek in order to provide written informed consent.

Once consent was acquired, the doctor used the form presented in Appendix A in order to log the observations needed to calculate VACS. Making the observations required to fill in the form did not interfere with the doctor’s work or alter the vaccination procedure in any way. In fact, the forms were actually filled in by the doctor after the patient’s visit was over, therefore the study did not affect the observed vaccination procedure in any way.

In addition to the VACS parameters, the form also records the doctor’s, parent/guardian’s and child’s own assessment of the vaccination experience. Doctors and parents/guardians are directly asked to distinguish between smooth, acceptable and bad procedures. Children’s feelings on the other hand are probed indirectly by asking whether they are willing to return to the doctor’s office for a future vaccination. Parents/guardians are also asked to report whether the child has had a prior bad vaccination experience or vaccination side effect.

The study’s protocol has been assessed and approved by the Research Ethics Committee of the University of Peloponnese (protocol code 20836/14.09.2022)

## 3. Results

### 3.1. Clinical data

Following the procedure described in section 2.4, vaccinations were carried out by 3 pediatricians in 1 public and 2 private pediatric clinics and their VACS parameters were recorded. The pediatricians submitted 45 records of vaccinations, of which 5 were discarded after a second check as the children involved were found to be outside the 2-12 years of age inclusion criterion (1 was a newborn, 4 were 13 years old or older). Thus 40 vaccination records were ultimately considered in this study.

The mean age of the participants was 8.79 years old, ranging from 2 years 0 months 4 days old to 12 years 6 months 25 days old. 25 of the participants were boys (62.5%) and 15 were girls (37.5%). 31 (77.5%) of the children had completed their recommended vaccination schedule while 9 (22.5%) of the children had skipped one or more of the vaccines recommended for their age group.

Figures 2, 3, 4 and 5 summarize the records for each of the observed parameters. To facilitate the reader in all graphs blue color has been assigned to the observation that does not indicate any discomfort, followed by yellow, orange and red in order of severity of the observed behavior. The most common discomfort indicator, observed in 19 (47.5%) of the cases, is lack of cooperation in stage III whilst the least common one is crying in stage I, observed in 5 (12.5%) of the cases.

**Figure 2.**
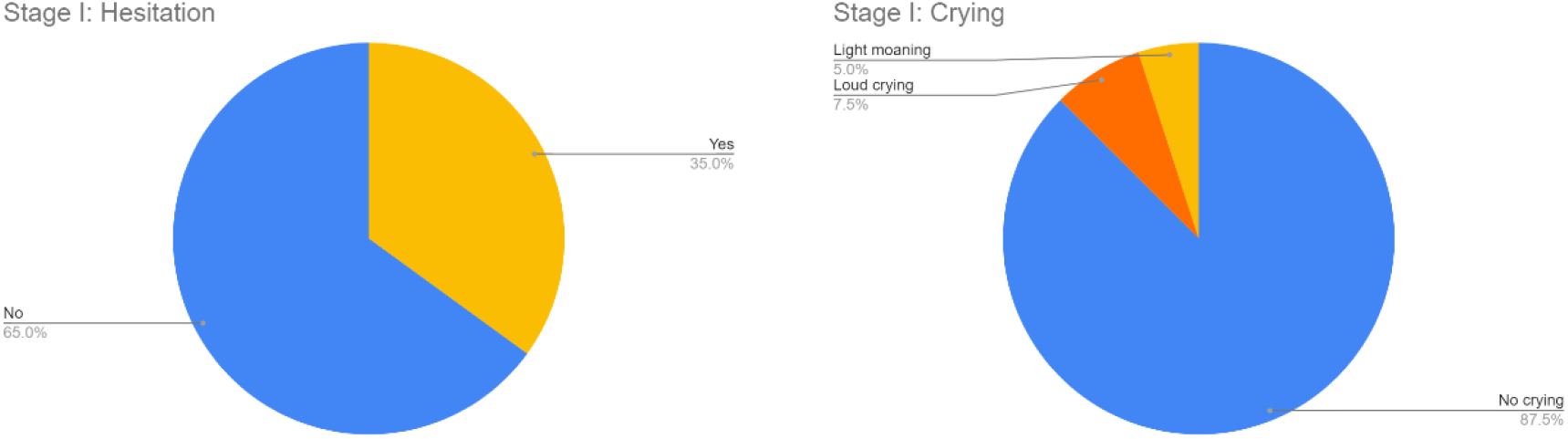
Stage I parameters. (a) Hesitation; (b) Crying.

**Figure 3.**
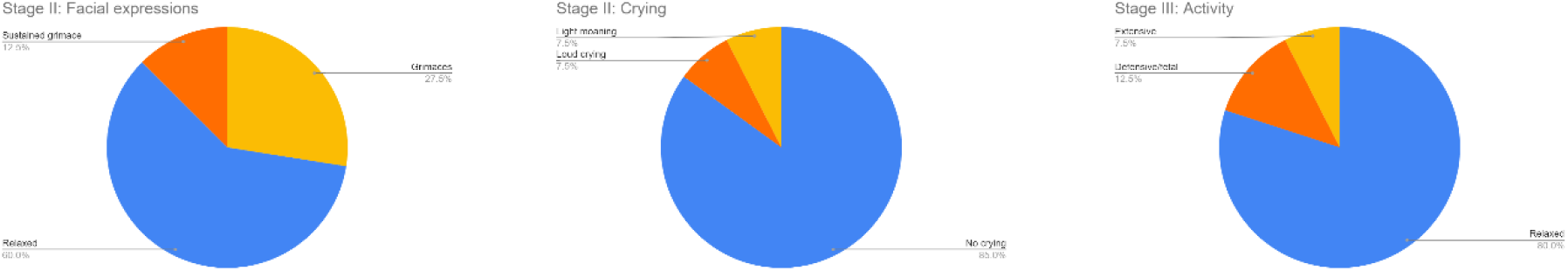
Stage II parameters. (a) Facial expressions; (b) Crying; (c) Activity.

**Figure 4.**
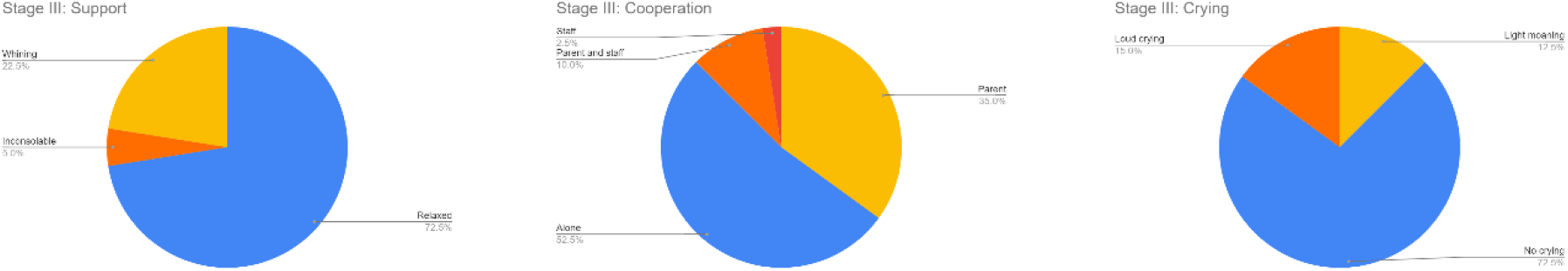
Stage III parameters. (a) Support; (b) Cooperation; (c) Crying.

**Figure 5.**
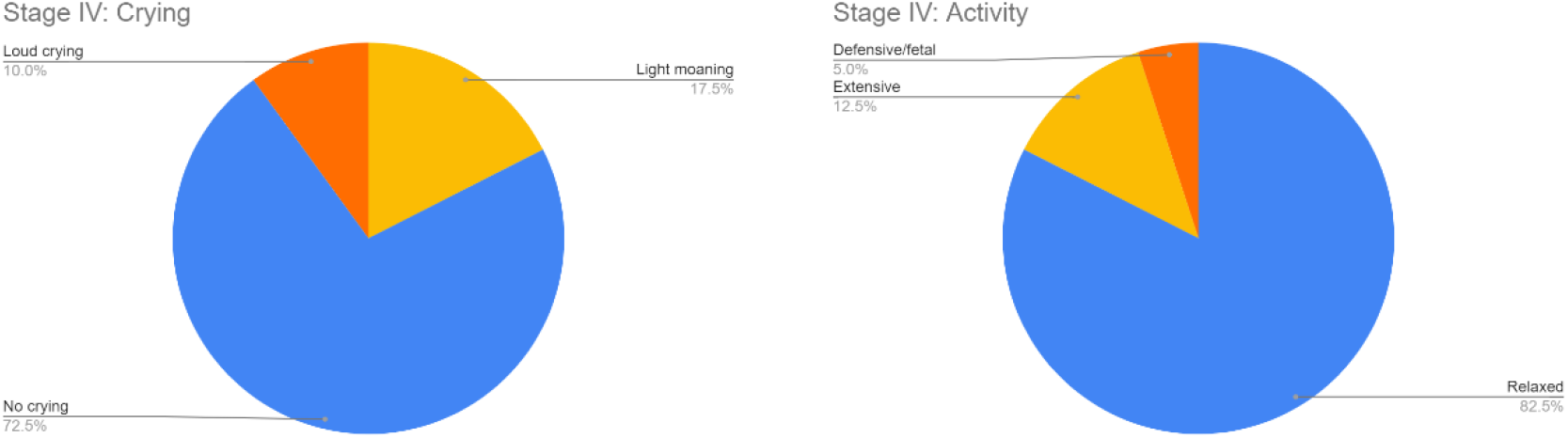
Stage IV parameters. (a) Crying; (b) Activity.

Doctors found the vaccination procedures to have run less than smoothly in 3 (7.5%) of the cases, while on the other hand children in 6 (15%) of the cases find the procedure so discomforting that they are reluctant to return for a future vaccination.

For 7 (17.5%) of the children their parents/guardians reported a prior bad vaccination experience. No prior vaccination side effects were reported.

All vaccinations were completed successfully, i.e. there were no cases in which the discomfort was so high that the vaccination had to be aborted.

### 3.2. VACS calculation and correlations

The weights presented in section 2.3 were used to calculate VACS for each of the 40 recorded vaccinations. A first finding is that vaccination discomfort is not a problem for all children, with 16 (40%) of the vaccinations having a VACS of exactly zero, i.e., the children did not display any indication of discomfort. The remaining children had VACS values ranging from 1 to 21 (median 6, mean 7.92, 95% CI:5.04-10.8). For the remaining of our study we have focused on the children that have non zero VACS, i.e., the children that have shown some discomfort during the vaccination procedure.

Doctors considered all vaccinations with a VACS number below 19 as smooth and all vaccinations with a VACS number above 19 as merely acceptable. This finding has two consequences:

1. VACS is validated as a measure of discomfort, with respect to doctors’ views: vaccination procedures that are deemed smooth by the doctors produce lower VACS values and less smooth procedures produce larger VACS values.
2. A threshold can be defined at VACS value 19, as the point above which vaccinations can no longer be deemed smooth.

Children, on the other hand, start stating that they are unwilling to return for a future vaccination when their experience has produced a VACS value as low as 11. This indicates that there is a disconnect between how children experience vaccination and how doctors consider children’s discomfort. It also suggests setting an even lower VACS threshold as a target: whilst a threshold of 19 is sufficient for doctors to consider vaccinations below the threshold as acceptable, it would take lowering the threshold to 11 to make sure that children are also content with their experiences.

One important question, of course, is that of predictability. If we can know be-forehand which children are in greater risk of experiencing discomfort, then perhaps we can focus more on how to best deal with them. Unfortunately, our data indicate that the parameters we have considered are poor predictors of VACS values.

Boys have an average VACS of 7.87 (95% CI:4.4-11.33) and girls an average VACS of 8 (95% CI:1.79-14.21). As also seen in Figure 6, there is no significant difference be-tween the genders.

**Figure 6.**
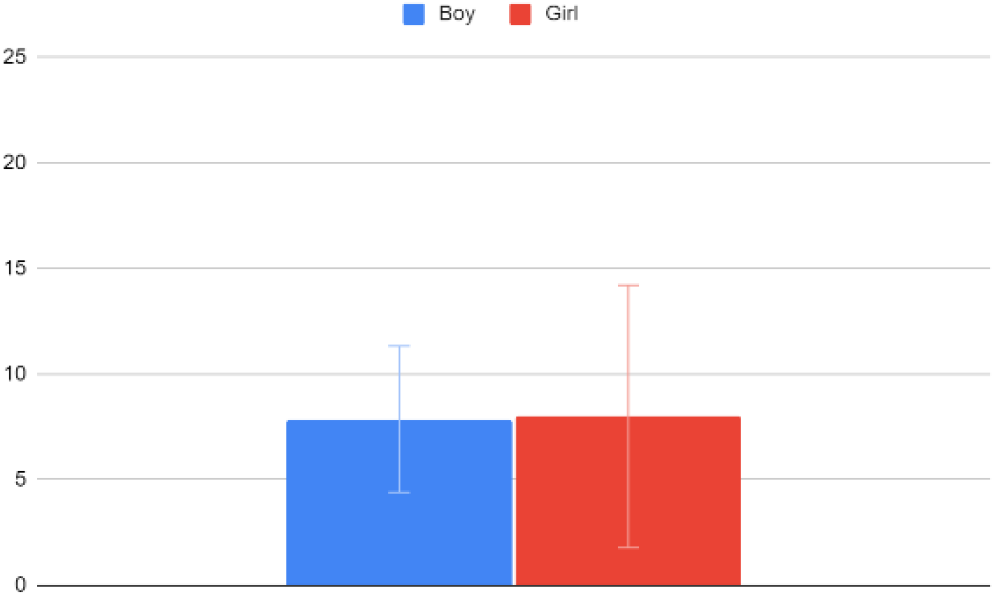
VACS values for boys and girls.

Children that have completed their recommended vaccination programs have an average VACS of 8.17 (95% CI:4.86-11.47) and children who have not completed their recommended vaccination programs have an average VACS of 7.17 (95% CI:-1.16-15.49). As also seen in Figure 7, there is no significant difference based on whether the children are fully vaccinated.

**Figure 7.**
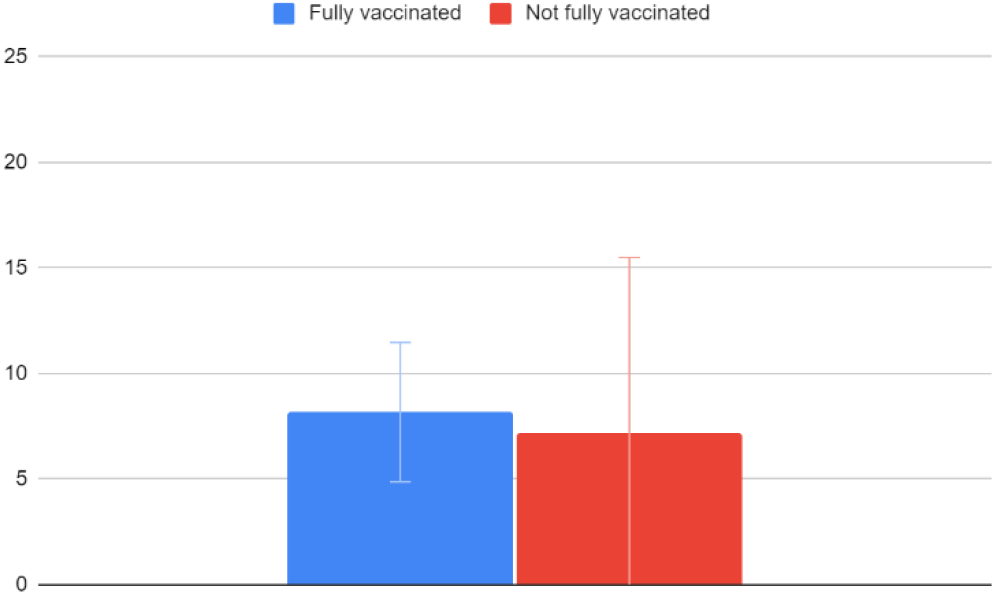
VACS values and vaccination history.

Children that have not had a previous negative vaccination experience have an average VACS of 7.41 (95% CI:3.78-11.04) and children who have had a bad experience before have an average VACS of 9.14 (95% CI:3.07-15.22). As also seen in Figure 8, although there is a small difference in the mean values, the confidence intervals are much wider making this difference of no significance.

**Figure 8.**
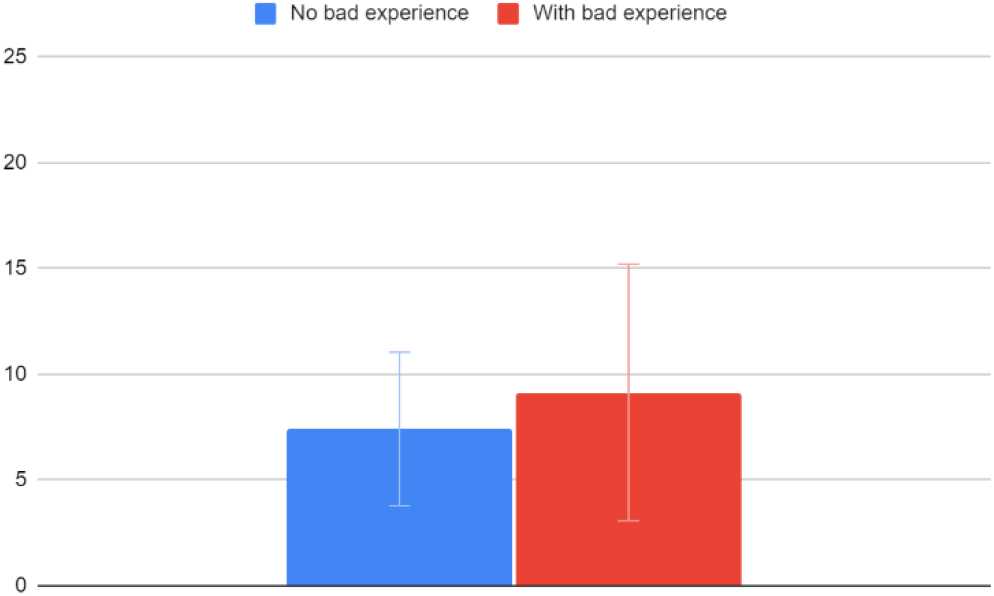
VACS values and bad vaccination experience history.

Children between the ages of 2-3 have an average VACS of 8.6 (95% CI:5.23-11.97), children between the ages of 4-7 have an average VACS of 9.75 (95% CI:2.41-17.09) and children between the ages of 8-12 have an average VACS of 6.27 (95% CI:2.6-9.94). Unsurprisingly, we find that older children tend to cope better with the vaccination procedure. Again, though, as also seen in figure 9, the confidence interval ranges are too broad to draw this conclusion safely.

**Figure 9.**
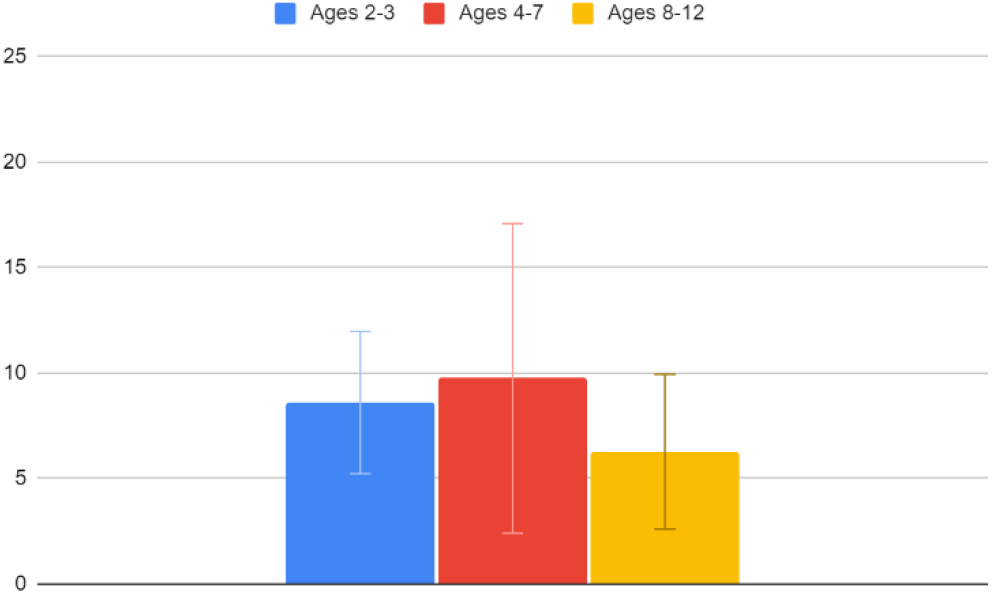
VACS values and age.

## 4. Discussion

There are many cases in which simple numerical scales are used in medicine to acquire quick and rough assessments of situations. These scales are not necessarily fully descriptive of all aspects of the observed phenomenon, but they are sufficient to provide a first idea of how close the situation is to the “normal” or “safe” range and an indication of whether some intervention is required or not. Examples include APGAR for newborns [1], Childhood Autism Rating Scale (CARS) for autism [2], Abbreviated Mental Test score (AMTS) for dementia [3], Montgomery–Åsberg Depression Rating Scale (MADRS) [4] for depressive episodes and so on. VACS aims to provide a similar tool for vaccination. It does not aim to measure discomfort to a great detail, but rather to provide a quick and rough estimation of how smoothly a vaccination has been for the child.

The medical scales that are most similar and relevant to VACS are those that aim to measure pain, in young children [5] and newborns [6]. But pain is not the only, and most probably not even the main source of discomfort for children during vaccination. This is evident by the fact that their discomfort often starts well before the actual vaccination, so it is clearly not induced by pain. A metric viewing discomfort from a broader perspective was needed; this is the gap that VACS aims to fill.

The fact that the majority of children display little or no discomfort is perhaps the reason that the alleviation of this discomfort has not been prioritized. We argue that this is not the medically prudent approach. As an exaggerated example, let us consider the case of poliomyelitis. Polio paralysis is much less frequent than severe vaccination discomfort (less than 1% of poliovirus infections result in paralysis [7] compared to 15% of the vaccinations resulting in such discomfort that the children are unwilling to be vaccinated again) but we consider polio as serious risk due to the severity of its complications in the rare occasions that they appear. In a some-what similar thinking, even if only a minority of children feel severe discomfort during vaccination, this discomfort is important to these children and it is our duty to seek ways to minimize it. Measuring this discomfort – instead of simply ignoring it and focusing only on completing the vaccination – is a first step in this direction.

### 4.1. Implications and future directions

The definition of VACS that is provided in this work can form the basis for further research and progress in the direction of comfortable vaccination. For example, having a tool with which to measure the discomfort experienced by children during vaccination, we can now use it in order to develop and then assess the effectiveness of alternative vaccination procedures. For example, a few years ago a VR system was presented for the discomfort free vaccination of children [8], but to this day no evidence has been provided regarding its effectiveness. With VACS, the efficacy of such solutions can now be quantified. Our team has already started to develop a proprietary VR system, similar to the one presented in [8], which we hope to assess using VACS and then release within the next couple of years.

Of course, high tech VR solutions are not necessarily the only way to improve children’s’ vaccination experiences. Other interventions in the vaccination procedure, ranging from the way the vaccination room is organized to the background music and from the timing of way the doctor talks to the child to the way the doctor is dressed, can now be examined and assessed.

The fact that the parameters examined in our study (gender, previous negative vaccination experiences and whether a recommended vaccination program has been completed) were found to be poor predictors of vaccination discomfort does not mean that vaccination discomfort is impossible to predict. Instead, it highlights the need for further research, with the examination of more parameters. This is an important research direction, as identifying the characteristics of children who experience intense vaccination discomfort may be key to understanding the core routes of this discomfort and designing mitigating measures.

Finally, individual pediatricians can use VACS to assess their own vaccination procedure and performance. If they find that their average VACS value to be high, perhaps there is reason to reconsider how they approach the vaccination procedure.

### 4.2. Weaknesses

Whilst the new scale is clearly validated (low values coincide with children’s, parents’ and doctors’ views that the vaccination was smooth and extreme discomfort produces the highest values), the small size of the observed sample does not allow for a very detailed estimation of the thresholds with high confidence. When a much larger number of vaccinations has been carried out and recorded, it will be possible to estimate more reliably the thresholds of VACS values for truly smooth, acceptable and unbearable vaccinations.

The information regarding previous negative experiences and previous vaccination side effects does not come from medical records but rather from the parents’ subjective reporting. Where one parent might consider an experience as ok and some side effects too minor to report, another parent might consider the same experience as negative and the same side effects as important. Therefore, the historical data regarding previous experiences and side effects are not equally reliable to the information observed by the doctors themselves.

Regarding the children’s opinion about their vaccination experience, it is more reliable for the older children. For children closer to the age of two, whose communication skills are still limited, their opinion is acquired with a much lesser certainty.

Finally, doctors’ assessments are also subjective as different doctors may assess the same observations differently. Where one doctor might record light moaning, an-other might note loud crying. As a result, VACS measurements are more reliably comparable only when they are performed by the same clinician; when different clinicians are involved, comparisons are safe only for larger differences between VACS measurements.

## Data Availability

All data produced in the present study are available upon reasonable request to the authors

## Author Contributions

Conceptualization, M.W.; methodology, M.W..; validation, M.W., S.A. and, V.P.; formal analysis, M.W. and V.P.; investigation, S.A.; resources, S.A.; data curation, M.W and V.P.; writing—original draft preparation, M.W.; writing—review and editing, V.P.; visualization, M.W and V.P.; supervision, M.W.; project administration, M.W.. All authors have read and agreed to the published version of the manuscript.

## Funding

This research received no external funding.

## Institutional Review Board Statement

The study was conducted in accordance with the Declaration of Helsinki and approved by the Research Ethics Committee of the University of Peloponnese (protocol code 20836/14.09.2022).

## Informed Consent Statement

Informed consent was obtained from the parents/guardians of all subjects involved in the study.

## Data Availability Statement

The data presented in this study are available upon reasonable re-quest from the corresponding author. The data are not publicly available due to their nature as they include personal records of individual minors.

## Acknowledgments

The authors express their gratitude to pediatricians K.M., B.C. and G.G. for their role in the application of VACS in a clinical setting.

## Conflicts of Interest

The authors declare no conflict of interest.

## Appendix

In the following images we present the form used to collect the data upon which the VACS calculation is based. We provide the English translation of the form, for international pediatricians who wish to incorporate VACS in their own clinical practice.

**Figure A2.**
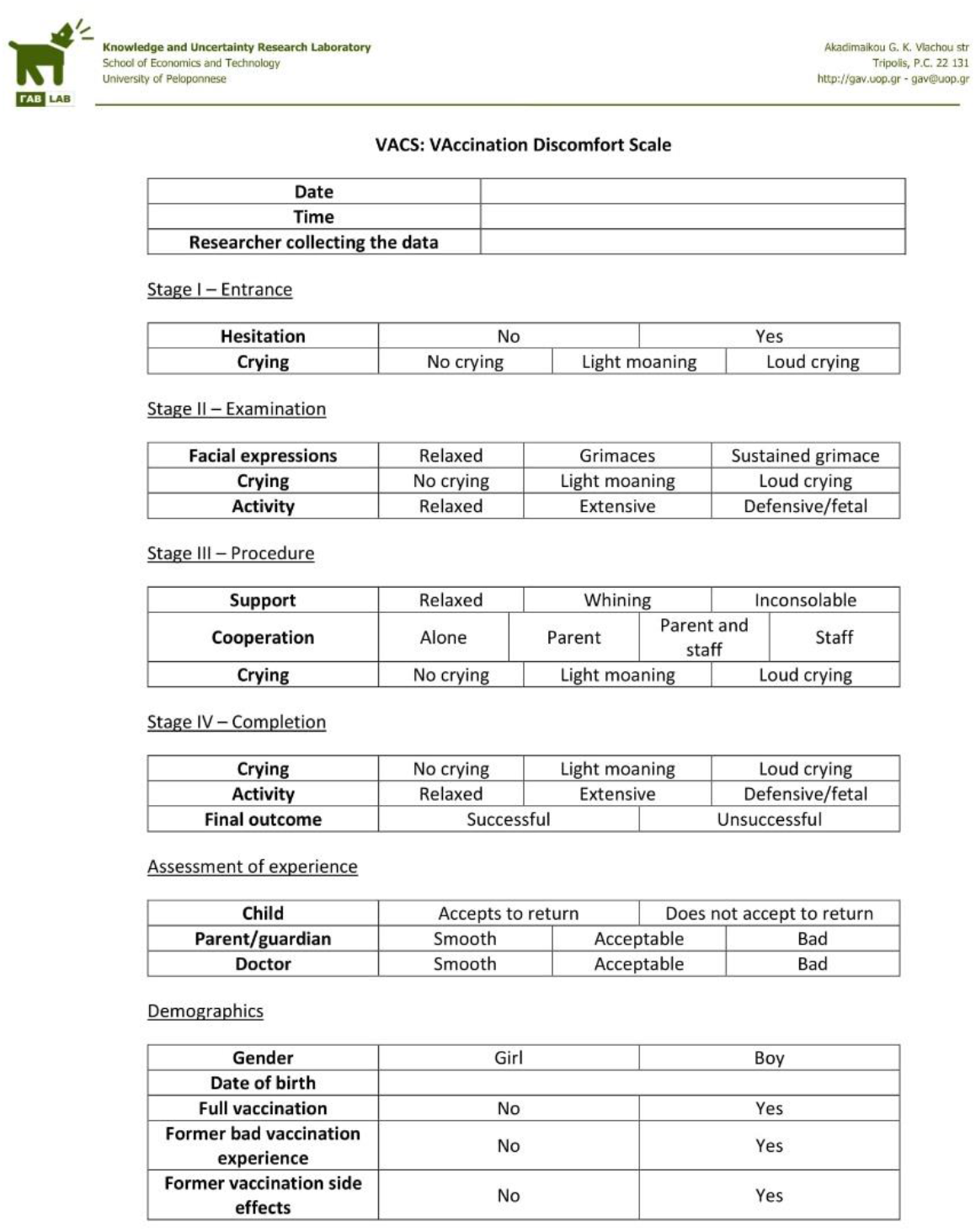
The form used to collect the data, translated into English.

## Notes

### Competing Interest Statement

The authors have declared no competing interest.

### Funding Statement

This study did not receive any funding

### Author Declarations

The Research Ethics Committee of the University of Peloponnese gave ethical approval for this work

## References

1. Apgar, V. A proposal for a new method of evaluation of the newborn infant. Curr Res Anesth Analg 1953, 32, 260–267.

2. Schopler, E.; Reichler, R.J.; DeVellis, R.F.; Daly, K. Toward objective classification of childhood autism: Childhood Autism Rating Scale (CARS). J Autism Dev Disord 1980, 10(1), 91–103. doi:10.1007/BF02408436

3. Hodkinson, H.M. Evaluation of a mental test score for assessment of mental impairment in the elderly. Age Ageing 1972, 1(4), 233–238. doi:10.1093/ageing/1.4.233

4. Montgomery, S.A.; Asberg, M. A new depression scale designed to be sensitive to change. Br J Psychiatry 1979, 134, 382–389. doi:10.1192/bjp.134.4.382

5. Merkel, S.I.; Voepel-Lewis, T.; Shayevitz, J.R.; Malviya, S. The FLACC: a behavioral scale for scoring postoperative pain in young children. Pediatr Nurs 1997, 23(3), 293–297.

6. Lawrence, J.; Alcock, D.; McGrath, P.; Kay, J.; MacMurray, S.B.; Dulberg, C. The development of a tool to assess neonatal pain. Neonatal Netw 1993, 12(6), 59–66.

7. Estivariz, C.F.; Link-Gelles, R.; Shimabukuro, T. Poliomyelitis. In Epidemiology and Prevention of Vaccine-Preventable Diseases, 14th ed.; Hall E., Wodi, A.P.; Hamborsky, J.; et al. Eds.; Publisher: Public Health Foundation, Washington USA, 2021, pp. 275–288.

8. VR Vaccine - Hermes Pardini | Our Work | Ogilvy. Available online: https://www.ogilvy.com/work/vr-vaccine (accessed on 16 October 2022).

